# PRState: Incorporating Genetic Ancestry in Prostate Cancer Risk scores for African American Men

**DOI:** 10.1101/2022.03.12.22271020

**Authors:** Meghana S. Pagadala, Joshua A. Linscott, James Talwar, Tyler Seibert, Brent Rose, Julie Lynch, Matthew Panizzon, Richard Hauger, Moritz H. Hansen, Jesse D. Sammon, Matthew H Hayn, Karim Kader, Hannah Carter, Stephen T. Ryan

## Abstract

Prostate cancer (PrCa) is one of the most genetically driven solid cancers with heritability estimates as high as 57%. African American men are at an increased risk of PrCa; however, current risk prediction models are based on European ancestry groups and may not be broadly applicable. In this study, we define an African ancestry group of 4,533 individuals to develop an African ancestry-specific PrCa polygenic risk score (PRState). We identified risk loci on chromosomes 3, 8, and 11 in the African ancestry group GWAS and constructed a polygenic risk score (PRS) from 10 African ancestry-specific PrCa risk SNPs, achieving an AUC of 0.61 [0.60-0.63] and 0.65 [0.64-0.67], when combined with age and family history. Performance dropped significantly when using ancestry-mismatched PRS models but remained comparable when using trans-ancestry models. Importantly, we validated the PRState score in the Million Veteran Program, demonstrating improved prediction of PrCa and metastatic PrCa in African American individuals. This study underscores the need for inclusion of individuals of African ancestry in gene variant discovery to optimize PRS.

## Introduction

Prostate cancer (PrCa) remains the most common non-skin malignancy in men, with significant mortality resulting in 1 in 42 men diagnosed with PrCa dying from the disease^1,2^. Men with an African ancestry have a 1.6 and 2.4 fold increased risk of PrCa diagnosis and age-matched mortality compared to men with a European ancestry^3,4^. Multiple studies suggest genetic heritability is high for PrCa^5,6^, with twin studies attributing 57% of PrCa risk to genetic factors^7^.

While rare high penetrance genes and missense mutations (e.g., G84E in HOXB13) have been described, they represent an exceedingly small minority of PrCa cases. Single nucleotide polymorphisms (SNPs) in non-coding regions also contribute to PrCa risk, with many falling in the chromosome 8q24 risk region^8,9^. Genome-wide association studies (GWAS) have identified more than 260 of these SNP susceptibility loci^10,11^. However, the majority of discovery populations in these GWAS have been of European or Asian ancestry and studies on the role of ancestral genetic background in PrCa risk for other ethnic groups are needed^8,10,12,13^.

Incorporating PrCa risk SNPs into a meaningful clinical tool is possible with polygenic risk scores (PRSs) that predict PrCa risk based on the presence of individual inherited SNPs^14–16^. Sun et al demonstrated that addition of a PRS to family history improved the performance of predicting PrCa in populations of predominantly European ancestry^11,16–20^. The full utility of such tools for diverse populations or in combination with nomograms is yet to be realized.

Here we use genetic ancestry to separate the ELLIPSE consortium into ancestry groups. We ran association studies within our African ancestry group to identify population-specific SNPs. We then constructed an African ancestry-specific PrCa PRS (PRState) that achieved an AUC of 0.65 [0.64-0.67] when combined with family history and age of diagnosis. Efficacy of PRS was contingent on inclusion of African ancestry group individuals in PRS construction. When only European ancestry group individuals were used to construct PRS, a considerable drop in performance was observed within the African ancestry group. PRS construction from trans-ancestry groups performed comparably to PRState.

Variants in PRState score have been described in previous trans-ancestry analysis; however, we demonstrate these variants contribute to a boost in PrCa prediction performance in both the ELLIPSE and Million Veteran Program African ancestry groups. These findings highlight the importance of ancestry-specific risk SNP identification and will hopefully guide future PRS studies of PrCa in African ancestry groups.

## Methods

### ELLIPSE Study Subjects and Genotype

The Elucidating Loci Involved in Prostate Cancer Susceptibility (ELLIPSE) consortium prostate cancer meta-analysis and genotypes (dbGaP Study Accession: phs001120.v1.p1) was accessed to analyze Affymetrix genotype calls for 91,644 male PrCa case/controls.

### Quality Assurance

PLINK (RRID:SCR_001757) genotype files consisting of 505,219 calls from the following consent groups were compiled: c1-c3,c6,c8,c10-18,c20,c23,c25,c27-28. Pre-imputation processing of autosomal and X chromosome genotypes followed below steps:

1. Duplicated variants were removed.
2. Heterozygous haploid SNPs were set to missing.
3. SNPs with call rate <90% were removed
4. SNPs with minor allele frequency (MAF) <1% were removed
5. Individuals with genotype coverage <90% were removed
6. Non-ACGT variants were removed.

Strand flips were reversed using snpflip. After preprocessing genotypes, the remaining 410,116 SNPs and 91,644 individuals were input to the secure Michigan Imputation Server (RRID:SCR_017579)^21^. Whole-genome SNPs were imputed with Minimac4 (RRID:SCR_009292) and ancestry-matched reference panel 1000 Genomes Project Phase 3 version 5 (RRID:SCR_008801). Finally, post-imputation duplicated SNPs and SNPs with MAF <1% were removed.

### Ancestry Likelihood Calculation (FROG-kb)

For ancestry group calculations, we opted for an ancestry group prediction tool that does not require relationships with other individuals, like PCA. FROG-kb^22^ uses Kidd AISNP panel (55 SNPs) to predict likelihood ratios for world geographic regions. Likelihood ratios for 160 populations were calculated and averaged. European American likelihood ratios were determined from populations in “Europe” region and African American likelihood ratios were determined from populations in “African” region. For the final European ancestry group, we used a European log likelihood > -10, resulting in 5567 individuals. For the final African ancestry group, we used a European log likelihood < -15 and African log likelihood > -15, resulting in 4533 individuals.

### Genome-wide association analyses (GWAS)

PLINK (RRID:SCR_001757) GLM method^23^ was used to conduct association analyses with PrCa case/control in European and African ancestry groups. All associations were adjusted for the first 10 principal components (PCA with 55-SNP Kidd panel) and age.

### Polygenic Risk Score Calculation

Association analyses within European, African or mixed ancestry training sets were conducted. Significant variants were identified through PLINK (RRID:SCR_001757) linkage-based clumping using a p1 threshold of 5e-08, a p2 threshold of 1e-05, an r2 threshold of 0.1 and a kb threshold of 1000 kb. Ten, seven and fourteen significant variants were identified through African, European and trans-ancestry analysis, respectively. For PRS construction, variants were weighted by log (base 10) odds ratio from the training set association statistics, oriented to PrCa risk allele, and combined. ROC-AUC evaluation across folds was conducted using polygenic scores as predictions.

For mismatched ancestry group analysis, European ancestry training sets were used for prediction on African ancestry test sets. For trans-ancestry group analysis, European and African ancestry training sets were combined and tested on African ancestry test sets. All three ancestry group PRSs were evaluated using 10-fold cross validation. AUC for each fold and overall are reported. Confidence intervals were calculated using pROC R package.

For contextualization of our results in relation to other genetic risk models, we compared our PRState score (composed of 10 African ancestry-specific variants) to variants published recently in a large meta-analysis of prostate cancer by Conti et al^11^. Half (5) of the variants used in the PRState score were in high linkage disequilibrium (r2 > 0.3) with Conti et al variants^11^. We excluded these variants and constructed a Conti PRS with reported odds ratio for the African group. To evaluate PRState and Conti polygenic risk scores, we used both scores as features for a logistic regression model with default parameters. Predicted probabilities were used in ROC evaluation. For the Million Veteran Program (MVP), genotype dosages were extracted for 10 PRState variants across participants and weighted by log (base 10) odds ratio from the best-performing fold in ELLIPSE. Performance was evaluated using ROC-AUC analysis in European and African ancestry groups.

Individual ancestry groups in the Million Veteran Program were characterized through Harmonized Ancestry and Race/Ethnicity (HARE) grouping^24^. HARE grouping was specifically developed to categorize MVP individuals based on self-reported ancestry and genetic ancestry. HARE utilizes a support vector machine to output probabilities of an individual’s ancestry group using self-identified and genetic ancestry. PrCa, metastatic PrCa and fatal PrCa status was determined through ICD 9/10 diagnosis, procedure code, CPT and HCPCS procedure code, laboratory values, medications and clinical notes from inpatient, outpatient and fee-based care in the VA healthcare system. Family history information was available for only 55,610 of 121,964 African individuals and 322,706 of 461,627 European individuals in MVP. When evaluating genetic information only, the full population was used and the subset of the population with family history information available was used for additional multivariable association. ROC-AUC analysis was conducted in European and African ancestry individuals separately.

## Results

### European and African Ancestry Group Identification in ELLIPSE

PCA analysis of 55 ancestry informative markers (AIMs) proposed by Kidd et al^25^ revealed that individuals in the ELLIPSE could be stratified according to ancestral background. European, African and Asian descent individuals formed distinct clusters **(Figure 1)**. European and African ancestry likelihood thresholds were selected such that population size was maximized while minimizing admixture. The European ancestry group included individuals with European ancestry likelihood ratio > -10, resulting in 5,567 individuals. The African ancestry group included individuals with African ancestry likelihood ratio > -15 and European ancestry Likelihood ratio < -15, resulting in 4,533 individuals **(Figure 1)**. PCA analysis confirmed thresholds results in tightly clustered European and African ancestry groups. Self-identified ancestry aligned with genetic ancestry defined through AIMs; although some individuals self-identifying as Hispanic were included in European and African ancestry groups (**Table 1)**.

**Table 1:**
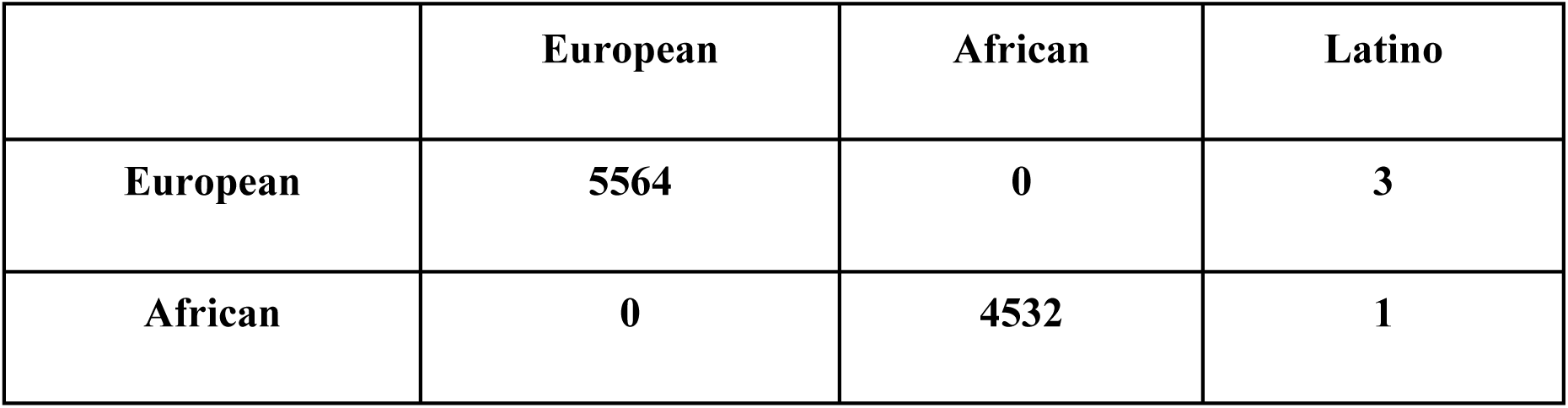
Self-Identified Ancestry of Genetically-Defined Ancestry Groups. Rows represent ancestry groups based on genetic ancestry. Columns represent self-identified ancestry. Values represent the number of individuals in each ancestry group and how they self-identify.

**Figure 1:**
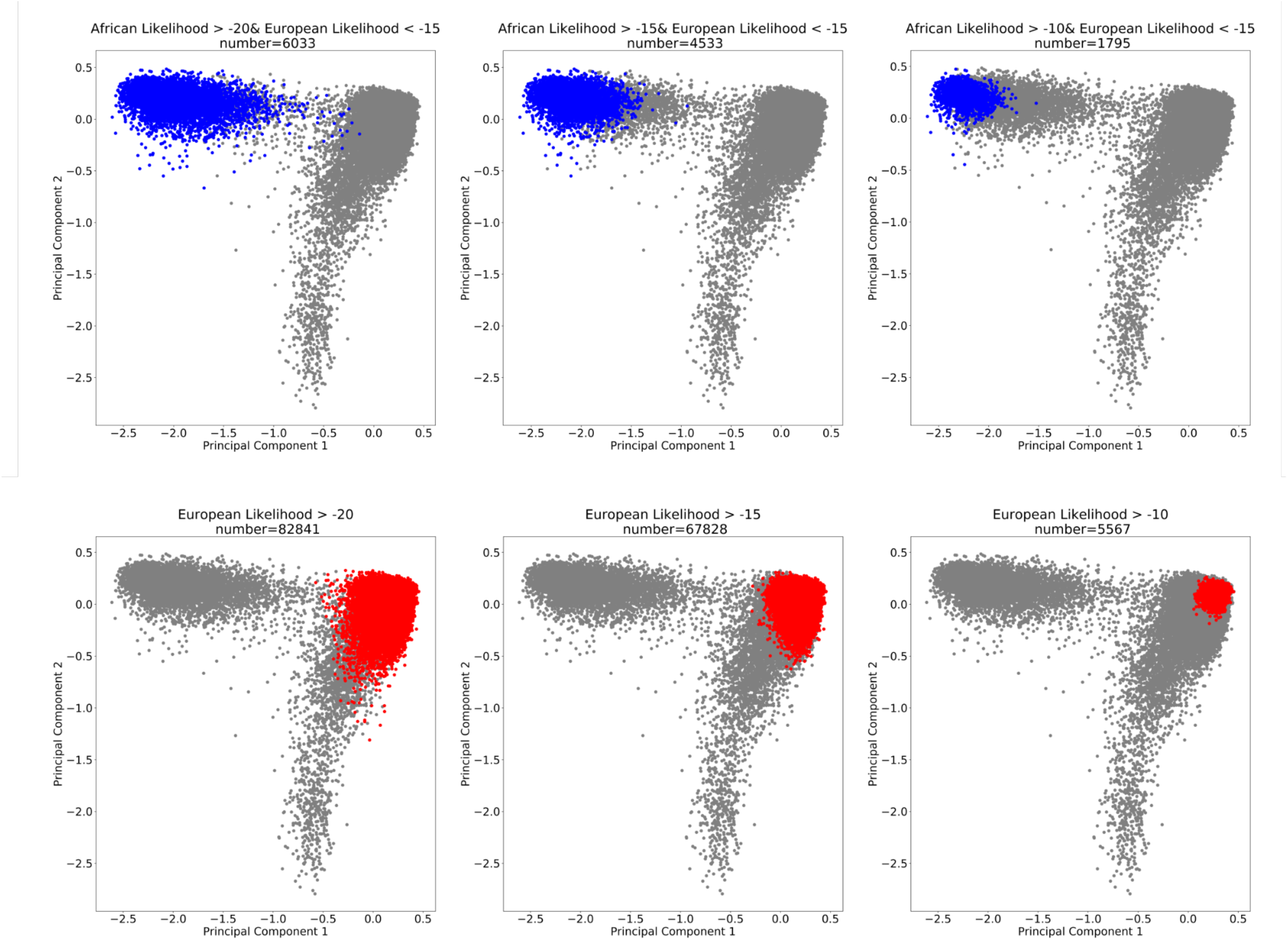
FROG-kb Ancestral Group selection. PCA analysis was conducted with Kidd et al. ancestry informative markers (AIMs). Identified groups were determined from European and African ancestry likelihood ratios (FROG-kb, Kidd et al. panel).

### Inclusion of African Ancestry Group Individuals in Polygenic Risk Score Construction Improves African Ancestry Group Prostate Cancer Prediction

GWAS was performed in the African ancestry group to identify African ancestry-specific risk loci. Loci on chromosomes 3, 8, and 11 were significantly associated with PrCa risk in African American men **(Figure 2, Supplementary Table 1)**. Age, family history, genetic risk score and the combination of these 3 factors for PrCa were evaluated using ROC-AUC analysis. Risk prediction models using genetics resulted in an AUC of 0.61 [0.60-0.63] **(Figure S1A)** while a combined model (genetics, age and family history) resulted in an AUC of 0.65 **(Figure 3A)**. To determine the efficacy of using a matched ancestry group model for prediction, we evaluated performance of a European ancestry group model on prediction of PrCa in our African ancestry group **(Supplementary Table 2)**. Interestingly, model performance was poor with near random performance using genetics [AUC: 0.52 (0.50-0.53)] (**Figure S1B)**. When genetics were combined with age and family history, the AUC was 0.59 [0.57-0.60], significantly lower than results from using the matched ancestry group model (**Figure 3B)**.

**Figure 2:**
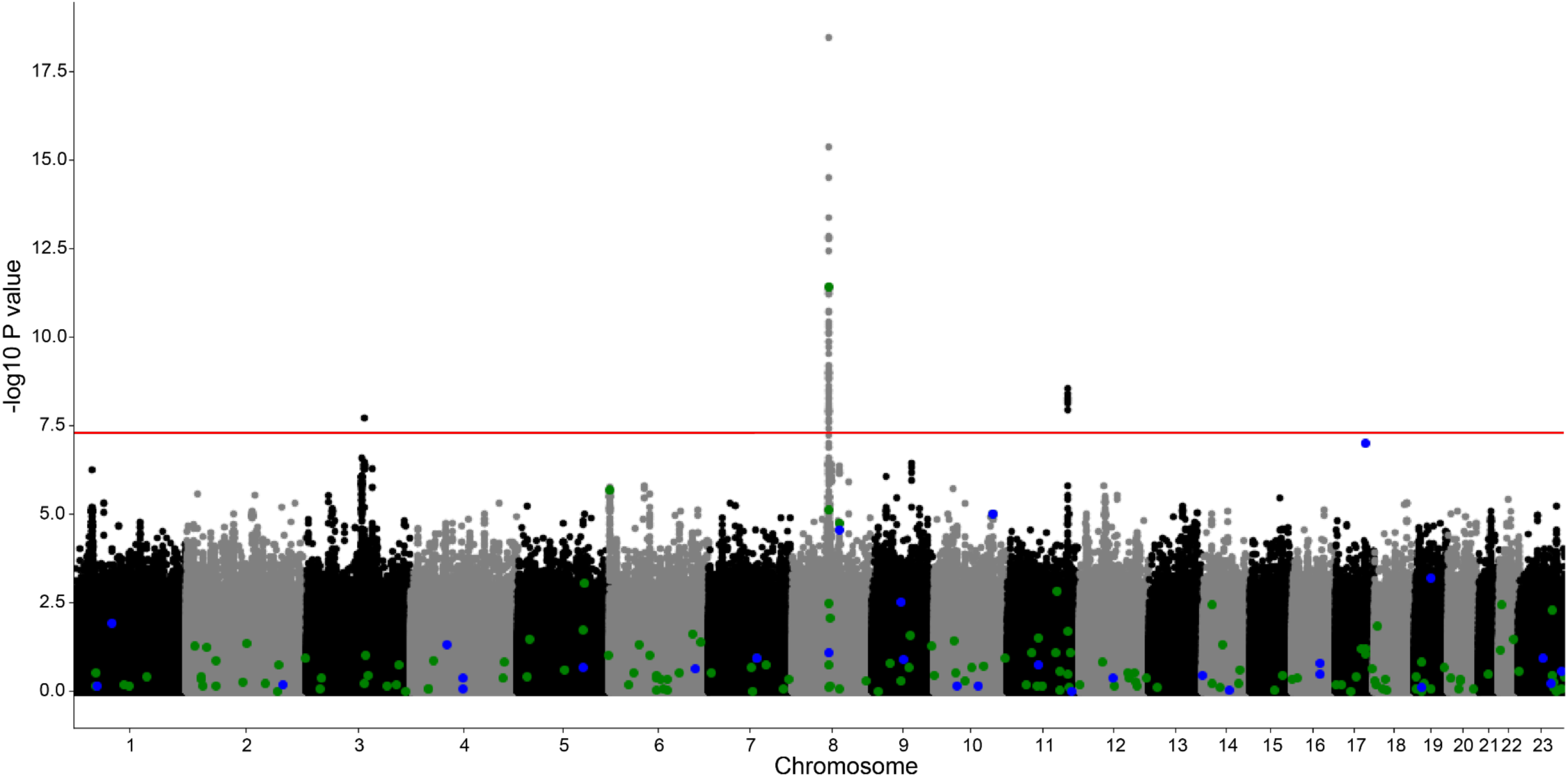
Manhattan Plot of Prostate Cancer (PrCa) Risk. Manhattan plot of logistic association of genetic variants with PrCa risk in ELLIPSE African ancestry group (n=4,533). green=known PrCa risk SNPs, blue=SNPs associated with PSA levels

**Figure 3:**
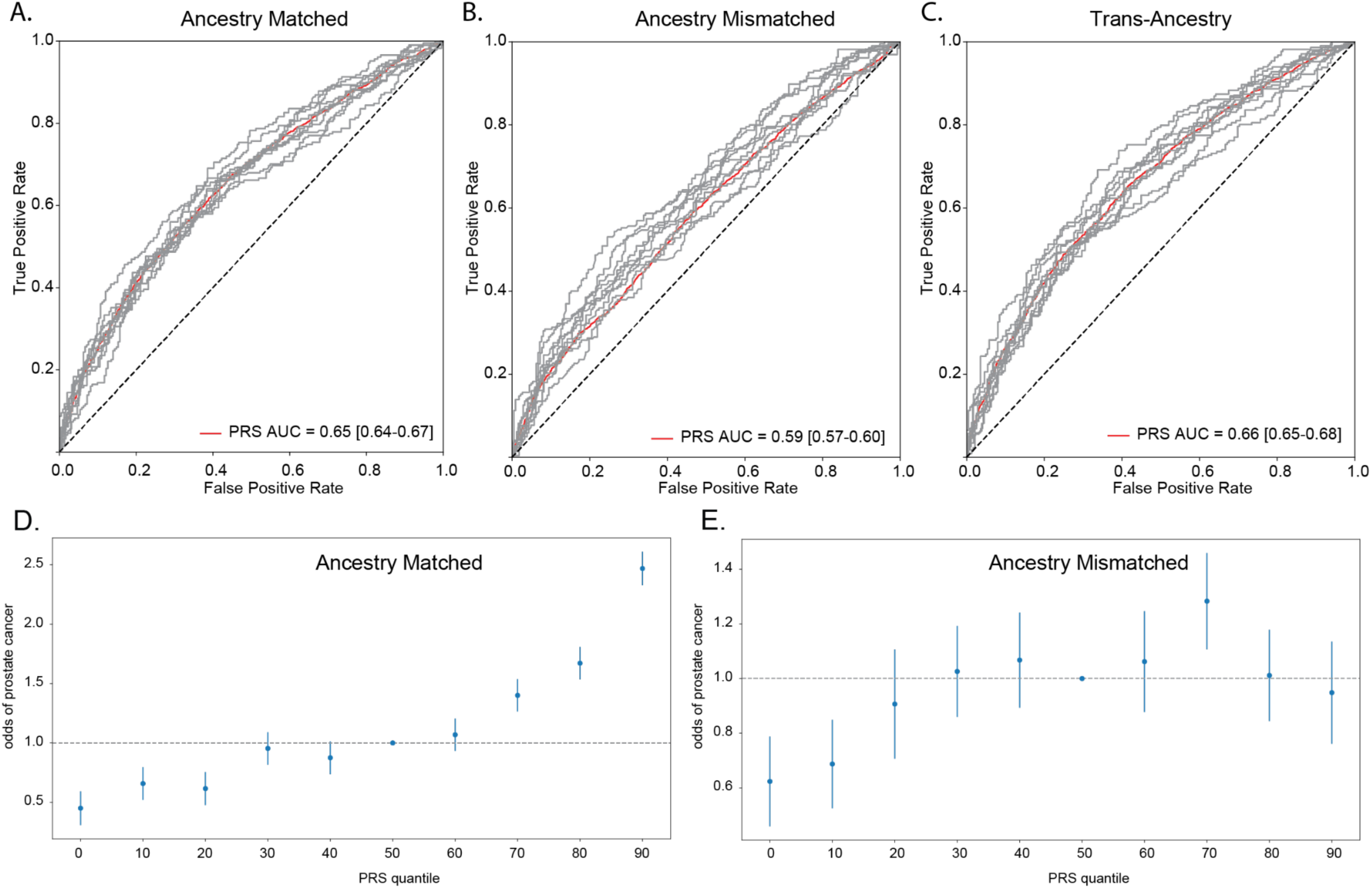
Performance of Polygenic Risk Scores (PRSs) Constructed from Different Ancestral Backgrounds in African Ancestry Group in ELLIPSE Consortium. ROC curve for genetic prediction of PrCa risk in ELLIPSE Consortium African ancestry group (n=4,533) using: **(A)** PRSs constructed from 10 African ancestry-specific variants with age and family history. **(B)** PRSs constructed from 7 European ancestry-specific variants with age and family history. **(C)** PRSs constructed from 14 trans-ancestry specific variants with age and family history. Quantile plot of PRS constructed from: 10 African ancestry-specific variants **(D)** and 7 European ancestry-specific variants **(E)** and respective odds of prostate cancer.

Lastly, recent studies of trans-ancestral analysis of PrCa risk have been conducted^11^, so we wanted to evaluate the performance of using a trans-ancestry group model on our African ancestry group. We trained models on a training set composed of both European and African ancestry groups combined and tested on the African ancestry group **(Supplementary Table 3)**. We achieved comparable performance to matched ancestry group models. We achieved an AUC of 0.62 [0.61-0.64] using only genetics **(Figure S1C)** and 0.66 [0.65-0.68] when combined with age and family history **(Figure 3C)**. Individuals in the top 10th quantile of PRS constructed from matched ancestry model had 2-fold greater risk of PrCa compared to the 50th quantile (**Figure 3D)**. Quantile analysis with odds of PrCa using a European ancestry group model demonstrates no trend between PRS and PrCa risk in the African American ELLIPSE Consortium (**Figure 3E)**.

Since the trans-ancestry group model used 5 variants which overlapped with the African ancestry group model, we compared odds ratios of African ancestry-specific risk variants in European, African and trans-ancestry groups to determine if variants exhibited different associations with PrCa **(Supplementary Table 4)**. Odds ratios between trans-ancestry group and African ancestry group association analyses were similar, compared to European ancestry group association analysis **(Figure S2)**. Odds ratios for African ancestry group individuals for 3 of the 8 chromosome 8 variants included in PRS construction were significantly different compared to European ancestry group individuals. These results demonstrate that PrCa variants have different effects based on ancestral background.

### African Ancestry PrCa Risk Variants Improve Prostate Cancer Prediction when Combined with Previous PrCa Risk Variants

After demonstrating the importance of inclusion of ancestry-matched individuals in polygenic risk score prediction, we sought to evaluate how our PRS constructed with 10 African ancestry-specific variants, which we will now refer to as PRState, performed in comparison to previous models. We compared performance to a PRS from a previously published large trans-ancestry analysis of PrCa by Conti et al.^11^. Five of the 10 African ancestry-specific variants we identified were in high linkage disequilibrium with variants previously implicated by Conti et al. and 2 of these 5 variants passed genome-wide significance threshold in Conti et al. African ancestry group GWAS. We compared our PRState score with the Conti et al. PRS constructed excluding these 5 variants. Addition of PRState score to Conti et al. PRS significantly improves prediction of PrCa by itself (DeLong P < .0003) (**Figure 4A)** and with family history and age (DeLong P < .0003) **(Figure 4B)**.

**Figure 4:**
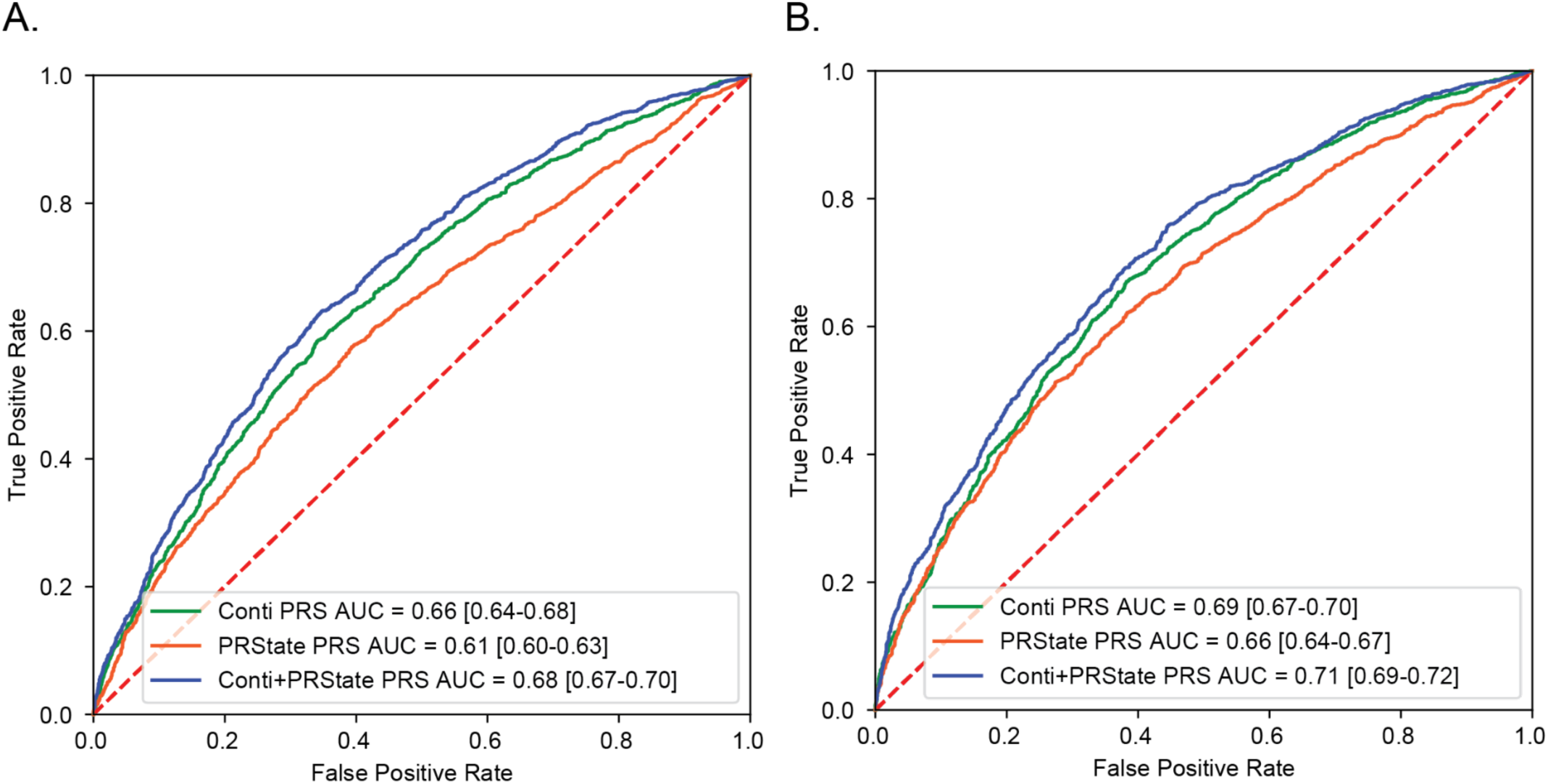
Evaluation of PRState and Conti Score in ELLIPSE Consortium. **(A)** ROC curve for genetic prediction of PrCa risk in ELLIPSE Consortium African ancestry group (n=4,533) using Conti PRS, PRState PRS and combined Conti and PRState PRS (DeLong p < 0.0003). **(B)** ROC curve for genetic prediction of PrCa risk in ELLIPSE Consortium African ancestry group (n=4,533) using Conti PRS, PRState PRS and combined Conti and PRState PRS along with family history and age (DeLong p < 0.0003).

To validate our results, we compared PRState score and Conti et al. PRS performance in the Million Veteran Program. Average AUC for PrCa **(Figure 5A)** (DeLong P < 1e-16) and metastatic PrCa **(Figure 5B)** (DeLong P < 8e-06) prediction was significantly higher in individuals of African ancestry when PRState and Conti et al PRS were combined compared to Conti et al. PRS alone. Combined PRState and Conti score was not associated with significantly higher AUC in predicting death from PrCa **(Figure 5C)** (DeLong P < .67). Interestingly, we find that PRState score performance was associated with significantly better predictive value compared to European individuals for all three defined clinical PrCa endpoints **(Figure 5D, E, F)** (PrCa DeLong P < 1e-16, metastatic PrCa DeLong P < 1e-16, fatal PrCa DeLong P < 1e-16). Furthermore, we characterized odds ratios of PRState variants between African and European HARE groups in the Million Veteran Program and noted a significant difference in the odds ratios for certain variants in all three endpoints (PrCa diagnosis, metastasis and death) (**Figure S3)**. These results not only demonstrate the need to include individuals of African ancestry in construction of polygenic risk scores that predict PrCa risk, but also that African ancestry-specific variants are critical for prediction of other PrCa characteristics, such as metastasis.

**Figure 5:**
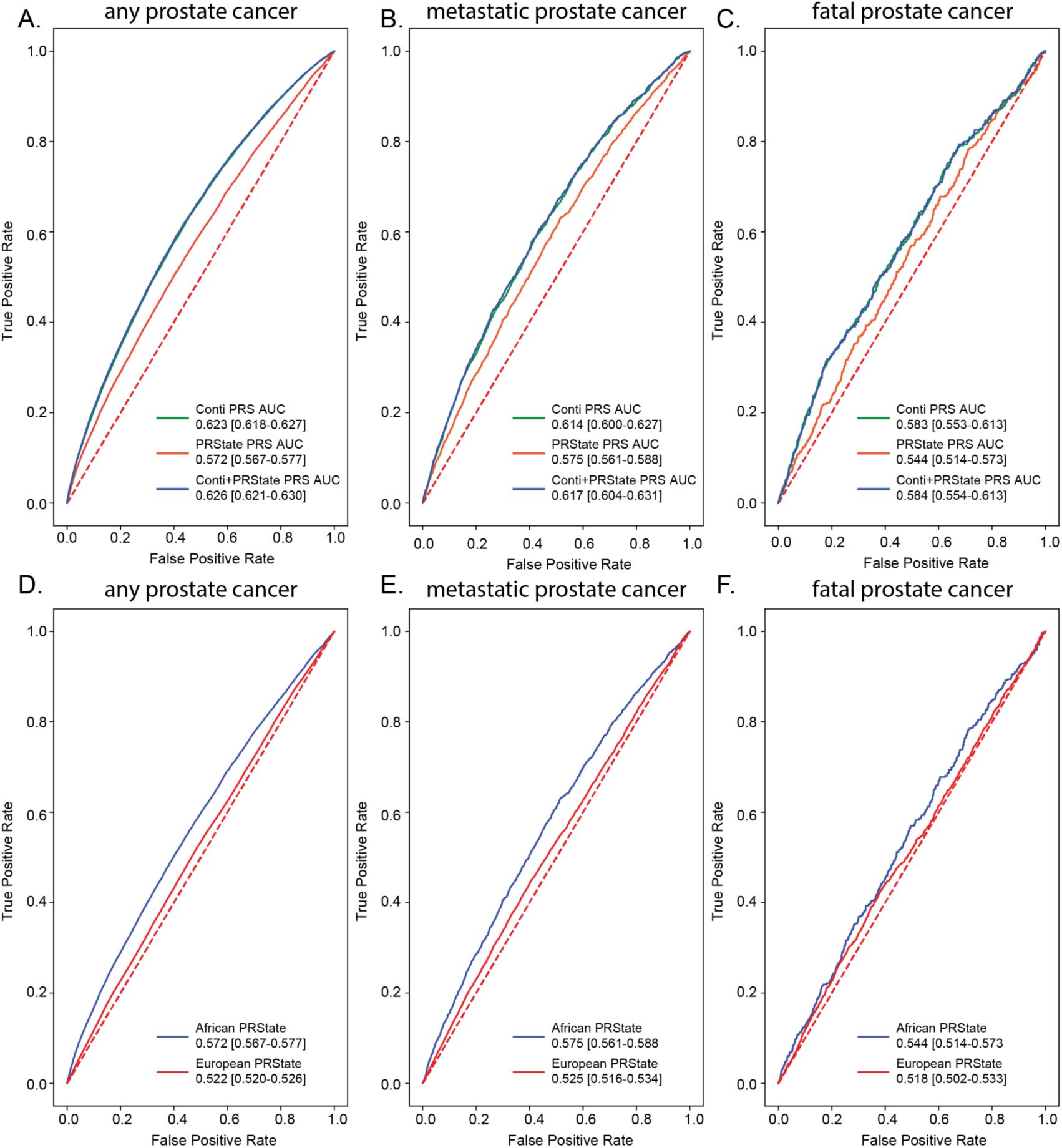
Evaluation of PRState and Conti Score in Million Veteran Program. ROC curve for genetic prediction of prostate cancer risk in Million Veteran Program African HARE group (n=121,964) using Conti PRS, PRState PRS and combined Conti and PRState PRS for any prostate cancer (**A)**, metastatic prostate cancer (**B)**, and fatal prostate cancer (**C)**. ROC curve for genetic prediction of prostate cancer risk in Million Veteran Program African HARE group (n=121,964) and European HARE group (n=461,627) using Conti PRS, PRState PRS and combined Conti and PRState PRS for any prostate cancer (**D)**, metastatic prostate cancer (**E)**, and fatal prostate cancer (**F)**.

## Discussion

A critical limitation in the majority of genetic studies in PrCa has been the overrepresentation of men with non-Hispanic European ancestry. Considering both the higher incidence and mortality of PrCa in African American men, this problem prevents the discovery of gene variants conferring PrCa risk in African and other ancestries. Using PrCa risk SNPs identified to be specific for African American men in ELLIPSE consortium, we used variants on chromosomes 3, 8, and 11 to construct an African ancestry-specific polygenic score (PRState) that achieved AUC of 0.61 [0.60-0.63] and 0.65 [0.64-0.67], when family history and age were added. We then compared PRState performance to models constructed from a primarily European ancestry group and a mixed European and African ancestry (trans-ancestry) group. We achieved comparable performance using a trans-ancestry group model, but a drop in performance using a European ancestry group model in ELLIPSE. The PRState score improved PrCa prediction performance when combined with a PRS constructed from a previous larger PrCa meta-analysis conducted by Conti et al.^11^ Although half of the PRState variants were in high linkage disequilibrium with Conti et al. variants, we demonstrate these African ancestry-specific variants significantly improve PrCa prediction in our discovery cohort (ELLIPSE) and external validation cohort (Million Veteran Program) of African ancestry group individuals. The results of our PRState study underscore the importance of including men with African ancestry when building genetic risk models and highlight African ancestry-specific PrCa variants that warrant further investigation.

PrCa is one of the most heritable cancer types and ancestry is an important determinant of PrCa risk. Using only a 55 SNP Panel and likelihood estimates from forensic genetic tool FROG-Kb, we were able to estimate genetic ancestry that aligned with self-identified ancestry in the ELLIPSE consortium. Although we had self-identified ancestry information available, our approach could be tested in cohorts where self-identified ancestry was not acquired. FROG-kb returns ancestry likelihood estimates for any panel of populations and individuals could have high ancestry likelihood estimates for several groups. For our study, we used FROB-kb to define categorical ancestry groups for PrCa risk variant discovery in the African ancestry group. Specifically, for defining the African ancestry group, we used a low European likelihood threshold combined with a high African likelihood threshold to define groups with little overlap in principal component analysis (**Figure 1)**. However, FROB-kb estimates do not have to be used categorically and can also be incorporated in the model. Specifically, for admixed individuals, this quantitative estimate of ancestry likelihood estimates may be useful in genetic risk models.

Despite a relatively small discovery cohort of 4,533 individuals, we were able to identify 10 African ancestry-specific variants (of which 5 were novel), that predicted PrCa in ELLIPSE similar to the PRS constructed from Conti et al. from over 250 variants (ELLIPSE PRState AUC = 0.66 [0.64-0.67] versus Conti PRS AUC = 0.69 [0.67-0.70]). Half of the PRState variants overlapped with variants identified by Conti et al., which included over 5x more individuals of African ancestry than our discovery cohort. Half of the PRState variants had been reported by Conti et al, whereas the novel variants included in PRState were identified using our methods. We applied a rigorous selection process for an African ancestry group that allowed identification of PrCa variants with African ancestry-specific effects. In order to test if our small sample was overfitting a prediction model, we applied our findings to an external cohort (the Million Veteran Program). PRState improved prediction of PrCa detection and demonstrated these variants were critical for prediction of high-risk PrCa that leads to metastasis in an African ancestry group within the Million Veteran Program. PRState was also associated with significantly higher AUC in the African ancestry group compared to European ancestry group for PrCa, metastatic PrCa and fatal PrCa. These results suggest PRState improves prediction of metastatic prostate cancer, which may help to prevent overtreatment of clinically non-aggressive prostate cancer with good survival

In the above analysis, we found PrCa risk SNPs in men ofAfrican descent are located at distinct loci that differ from PrCa risk loci identified in men of European ancestry. Variants on chromosome 8 were identified in both European and African ancestry group PrCa GWAS, however, the African ancestry-specific chromosome 8 variants (rs113343238, rs16902008, rs943270004, rs116845582, rs59825493) were not significantly associated with PrCa risk in the European ancestry group in the ELLIPSE Consortium^26,27^. Interestingly, in the Million Veteran Program, certain variants were protective in the European ancestry group but associated with higher PrCa diagnosis in the African ancestry group (rs113343238, rs943270004). These results suggest that even within well-known PrCa risk loci, defining ancestry group differences will likely improve genetic risk models.

Our PRState analyses identified 10 African ancestry-specific variants in our discovery cohort of 4,533 individuals and demonstrated its improved predictive power. With a larger cohort, we could apply our approach to identify potential novel African ancestry-specific PrCa risk variants. While this work has demonstrated the feasibility of using a small number of SNPs to define ancestry backgrounds and then predict genetic risk of PrCa, there are several limitations. The ELLIPSE data set is the largest complete PrCa cohort and we focused on 4,533 patients in an African ancestry group representing <5% of the total ELLIPSE cohort. Thus the study was not powered to identify potentially meaningful SNPs, as indicated by the suggestive peaks in the African ancestry group GWAS on chromosomes 9, and 12, which did not reach statistical significance. Additionally, we recognize African ancestry encompasses a wide breadth of genetic diversity that can not be wholly defined by a small sample. Nevertheless, we believe our PRState study indicates the inclusion of ancestral inherited risk is an important variable for analyzing PrCa risk similar to family history. Further investigation of a larger African ancestry sample will likely improve the signal and determine the magnitude of PrCa risk conferred by the African ancestry-specific SNPs identified in our study. Additionally, while one unintentional risk of PrCa treatment is overtreatment of low grade PrCa disease, the ELLIPSE data does not include high grade versus low grade cancer characterization. Therefore, we are unable to identify whether these SNPs confer higher risk for PrCa metastasis and death, although our analysis within the MVP demonstrates an association. Future studies will expand these methods to other under-represented ancestry groups with a goal of developing PrCa risk stratifying tools based on an individual’s ancestral background. Increasing the number of non-white patients in databases such as the ELLIPSE consortium is a key element to furthering research in these groups.

## Conclusions

Prostate cancer demonstrates the highest genetic risk of all cancers, and sorting patients by genetic ancestry compared to self-identified ancestry allows discovery of heritable risk SNPs in men with different ancestries. We have shown that a 55 SNP panel can be used to separate European and African ancestral groups and to support identification of risk SNPs unique to African ancestry, which importantly improves prediction of PrCa risk in men of African descent, especially when combined with age and family history.

## Data Availability

Summary statistics are within the manuscript and Supporting Information files. The data analyzed in this study were obtained from dbgap (accession id: phs001120.v1.p1) and Million Veteran Program, Office of Research and Development, Veterans Health Administration. Access to raw genotype and phenotype data can be obtained through application.

## Acknowledgements

This research used data from the Million Veteran Program, Office of Research and Development, Veterans Health Administration. This research was supported by the Million Veteran Program MVP022 award #I01 CX001727 (PI: Richard L. Hauger MD). This publication does not represent the views of the Department of Veterans Affairs or the United States Government. This work was supported by National Institutes of Health grants #1F30CA247168, #T32CA067754 to Meghana S. Pagadala and an Emerging Leader Award from The Mark Foundation for Cancer Research grant #18-022-ELA to Hannah Carter.

## Ethics Statement

The Central Veterans Affairs Institutional Review Board (IRB) and site-specific Research and Development Committees approved the Million Veteran Program study. Analysis of ELLIPSE consortium analysis was approved through dbGaP. All relevant ethical regulations were followed.

## Author Contributions

S.T.R., H.C., J.A.L. and M.S.P conceived the work; M.S.P and J.A.L. conducted formal analysis and investigation; M.S.P. and J.A.L. wrote the paper with assistance from J.T., T.M.S., B.R., J.L., M.P., R.H., M.H.H., J.D.S., M.H.H., K.K., H.C. and S.T.R.; T.M.S., B.R., J.L. M.P. and R.H. assisted in MVP validation

## Supplementary Information

**Supplemental Table 1** ELLIPSE African Ancestry GWAS Statistics (n=4553)

**Supplemental Table 2** ELLIPSE European Ancestry GWAS Statistics (n=5567)

**Supplemental Table 3** ELLIPSE European and African Ancestry GWAS Statistics (n=10100)

**Supplemental Table 4** Significant Variants Identified in ELLIPSE European, African, and Trans-Ancestry Groups

## Code Availability Statement

Code used for this manuscript are at: https://github.com/meghanasp21/PRState

## Notes

### Competing Interest Statement

The authors have declared no competing interest.

### Author Declarations

The data analyzed in this study were obtained from dbgap (accession id: phs001120.v1.p1) and Million Veteran Program, Office of Research and Development, Veterans Health Administration. Access to raw genotype and phenotype data can be obtained through application.

