## Supplemental Figures for "PRState: Incorporating Genetic Ancestry in Prostate Cancer Risk scores for African American Men"

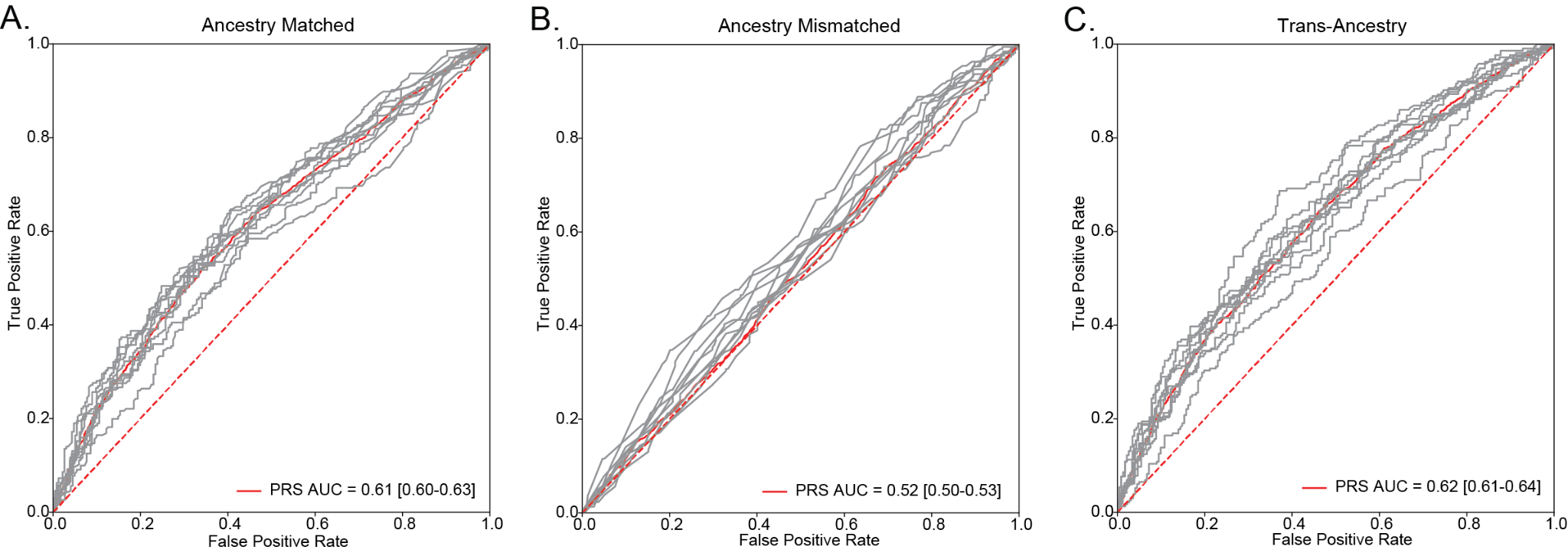

**Supplemental Figure 1: Performance of Polygenic Risk Scores Constructed from Different Ancestral Backgrounds in African Ancestry Group in ELLIPSE Consortium.** ROC curve for genetic prediction of prostate cancer risk in ELLIPSE Consortium African ancestry group (n=4,533) using: **(A)** Polygenic risk scores constructed from 10 African ancestry-specific variants only. **(B)** Polygenic risk scores constructed from 7 European ancestry-specific variants only. **(C)** Polygenic risk scores constructed from 14 trans-ancestry specific variants only.

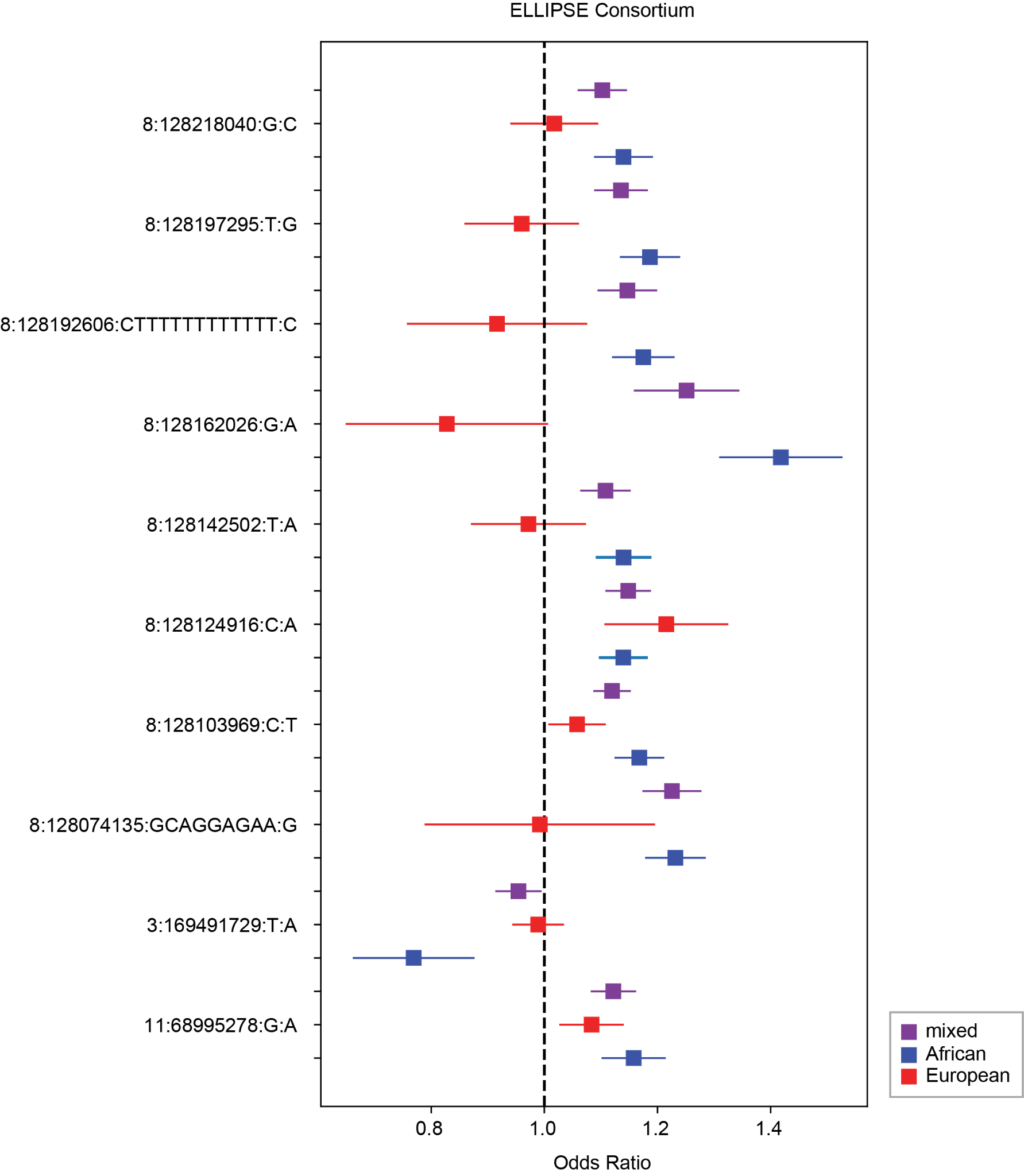

**Supplemental Figure 2: Comparison of African Prostate Cancer Variant Odds Ratios in European, African, and Trans-Ancestry Groups in ELLIPSE Consortium.** Odds ratios for 10 African ancestry-specific PrCa variants for prostate cancer in the ELLIPSE Consortium for European (n=5,667), African (n=4,553) and Trans-Ancestry (n=10,100) ancestry groups.

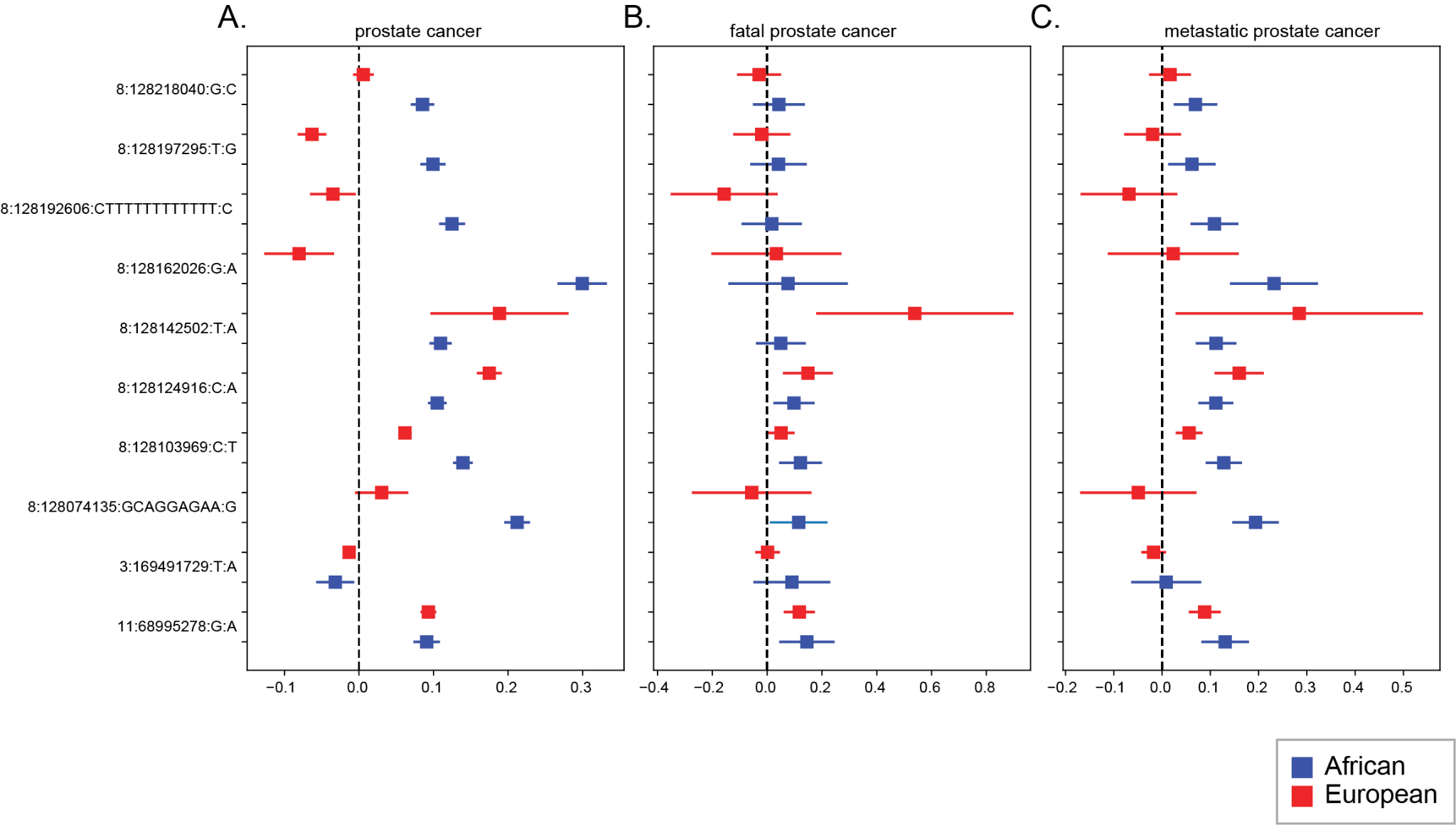

**Supplemental Figure 3: Comparison of African Prostate Cancer Variant Odds Ratios in European and African HARE Groups in Million Veteran Program.** Odds ratios for 10 African ancestry-specific PrCa variants for any prostate cancer **(A),** fatal prostate cancer **(B),** and metastatic prostate cancer **(C)** in the African (n=121,964) and European (n=461,627) HARE Million Veteran Program groups.
